# Sex-Based Differences in Clinical Presentation, Management, and Outcomes of Acute Coronary Syndrome in Brazilian Emergency Medical Services

**DOI:** 10.64898/2026.08.17.26360642

**Authors:** Antonio Fagundes, Andrea D. Stephanus, Renata Moll-Bernardes, Denilson Albuquerque, Angelina Silva Camiletti, Emiliano Horacio Medei, Andre Feldman, Marcia M Noya-Rabelo, Rose Mary Frajtag, Olga Ferreira de Souza

## Abstract

**Background:** Sex-related disparities in acute coronary syndrome (ACS) recognition and management remain an important global health concern. Women frequently present with less common symptoms and may experience delays in diagnostic evaluation. This study examined sex-based differences in clinical presentation, management, and outcomes among patients with acute chest pain attended by emergency medical services (EMS) across Brazil.

**Methods:** We conducted a retrospective multicenter study using a national registry database from 14 Brazilian states between January 2020 and June 2024, within a large private hospital network. All patients presenting with acute chest pain were classified by trained cardiologists as unstable angina (UA), ST-elevation myocardial infarction (STEMI), or non–ST-elevation myocardial infarction (NSTEMI). Multivariable regression models, with men as the reference group, evaluated sex differences in diagnosis, treatment, and outcomes; categorical outcomes were modeled with logistic or multinomial logistic regression and continuous outcomes with linear regression. Sensitivity analyses included state-clustered standard errors and E-value quantification of potential unmeasured confounding.

**Results:** Among 7,171 patients with confirmed ACS (68.2% male), the median age was 63.0 years [IQR 20.0]; women were older than men (67.0 [20.0] vs 61.0 [19.0] years). Final diagnoses were UA in 46.7%, STEMI in 18.8%, and NSTEMI in 34.6% of patients. Men accounted for 75.5% of STEMI and 69.7% of NSTEMI cases. Overall, 91.7% received aspirin and 89.6% received at least one additional antiplatelet agent. After multivariable adjustment, women had higher odds of chest pain classified as probably ischemic and possibly ischemic compared with definitely ischemic chest pain (adjusted OR 1.51 [95% CI 1.33–1.72] and 1.60 [1.37–1.86], respectively) and lower odds of STEMI and NSTEMI relative to unstable angina (adjusted OR 0.59 [0.51–0.68] and 0.74 [0.66–0.83], respectively). Door-to-ECG time was longer in women in the unadjusted analysis (β=1.53 minutes [0.24–2.82]) but the difference was no longer significant after adjustment (β=1.04 minutes [−0.27 to 2.36]). In-hospital mortality did not differ between sexes in unadjusted analysis, and there was no evidence of excess short-term mortality in women.

**Conclusions:** Within a large private hospital network in Brazil, women with confirmed ACS were more often classified with less definitely ischemic type of chest pain and were less frequently classified as STEMI or NSTEMI than men. Differences in door-to-ECG time did not persist after adjustment, and in-hospital mortality did not differ by sex. These findings highlight the relevance of sex-sensitive triage and diagnostic protocols within EMS systems to reduce inequities in ACS recognition and treatment.

## Introduction

Acute coronary syndrome (ACS) remains a leading cause of morbidity and mortality worldwide.^1^ It continues to impose a substantial burden on emergency medical systems, particularly in low- and middle-income countries.^2^ Despite advances in diagnostic strategies and evidence-based therapies, sex-related disparities persist across the ACS care continuum.^3^ Women are more likely than men to present with different or less clearly ischemic symptoms, experience delays in diagnostic evaluation, and, in some settings, receive fewer guideline-recommended therapies. These inequities may translate into worse short- and long-term outcomes.^4^

Emergency medical services (EMS) represent a critical link in the ACS care chain. Early recognition of symptoms, timely electrocardiographic (ECG) acquisition, and rapid initiation of appropriate reperfusion pathways are central determinants of outcomes, particularly in STEMI.^5^ However, evidence from low- and middle-income countries is limited, suggesting similar or greater disparities.^6^

Brazil has an extensive, heterogeneous EMS network that operates under both public and private principles, with substantial variation in infrastructure, staffing, and local protocols.^7^

We therefore aimed to evaluate sex-based differences in clinical presentation, diagnostic timeliness, management, and outcomes among patients with ACS attended by EMS using a protocolized chest pain model in 14 Brazilian states. We hypothesized that women would present with different symptom profiles but would experience the same time-sensitive care and similar outcomes to men, due to a unified chest pain protocol.

## Methods

### Study Design and Setting

We conducted a retrospective, multicenter observational study using a national chest pain registry from EMS systems operating in 14 Brazilian states, within the largest network of private hospitals in Brazil, between January 1, 2020, and June 30, 2024. These services follow a unified chest pain protocol but differ in organizational structure, resource availability, and local practice patterns, providing a broad view of hospital ACS management across diverse regions. All EMS had cardiologists trained in the AHA/ACC guidelines for the management of patients with chest pain and ACS.^5,8^

The study was approved by the institutional review board of Daher Hospital under protocol number 8.559.285, and the requirement for informed consent was waived due to the retrospective use of de-identified registry data.

### Study Population and Stratification

We analyzed the national chest pain registry, which included all adult patients (≥18 years) who presented to EMS with acute chest pain during the study period and for whom subsequent ACS was confirmed as a final diagnosis. All data from EMS encounters were linked to hospital-level data on diagnoses, procedures, and outcomes. Final diagnoses were categorized as unstable angina (UA), ST-elevation myocardial infarction (STEMI), or non–ST–elevation myocardial infarction (NSTEMI). Patients with missing sex information were excluded. The study population was stratified by sex (female vs male) and by final diagnosis (UA, STEMI, NSTEMI) for descriptive and regression analyses. Because the cohort was restricted to patients with ACS confirmed as the final diagnosis, the analyses focus on sex-related differences in classification and management among patients already identified as having ACS, rather than on differences in recognition at first medical contact.

### Data Sources and Variables

Data were obtained from a national chest pain registry database. Collected variables included demographics, cardiovascular risk factors (hypertension, smoking, obesity, diabetes mellitus, dyslipidemia, sedentary lifestyle, and family history of premature coronary artery disease); chest pain category (definitely ischemic, probably ischemic, possibly ischemic, non-ischemic, anginal equivalent, and evolved myocardial infarction); aspirin administration, other antiplatelet administration, non-invasive functional testing and imaging; cardiac catheterization; number of coronary arteries treated; final diagnosis (unstable angina, STEMI and NSTEMI); time metrics (door-to-ECG time, door-to-needle time, and door-to-guidewire time); and outcomes (closed-unit length of stay, hospital length of stay, and in-hospital mortality).

Chest pain category was assigned by the attending cardiologist according to the registry protocol and therefore reflects the clinician’s classification rather than patient-reported symptoms. Time metrics were measured from hospital arrival. Door-to-needle and door-to-guidewire times were evaluated only in patients with STEMI undergoing fibrinolysis or percutaneous coronary intervention, respectively.

### Handling Missing Values

Missing values were addressed using multiple imputation via the mice (Multivariate Imputation by Chained Equations) package in R. Imputation models were tailored to the variable type: predictive mean matching for continuous variables, logistic regression for binary variables, and polytomous logistic regression for categorical variables with two or more levels. The plausibility of imputed values, residual distributions, and convergence diagnostics were assessed.

Baseline covariates with missing data were imputed, whereas outcome variables were not imputed to avoid introducing bias. Five imputed datasets were generated and combined using Rubin’s rules for descriptive analyses and regression modeling. Because outcomes were not imputed, the number of observations varied according to outcome availability.

### Outcomes and Covariates

The outcomes comprised three domains: diagnostic classification (chest pain category and final ACS diagnosis), management (aspirin administration, other antiplatelet use, non-invasive stratification, cardiac catheterization, and number of coronary arteries treated), and in-hospital course (closed-unit length of stay, hospital length of stay, door-to-ECG time, and in-hospital mortality).

Covariates for multivariable adjustment included age, hypertension, smoking, obesity, diabetes mellitus, dyslipidemia, sedentary lifestyle, and family history of premature coronary artery disease.

### Statistical Analysis

Continuous variables were summarized as mean ± standard deviation (SD) or median [interquartile range (IQR)], as appropriate, and categorical variables as counts and percentages. Normality was assessed with histograms, normal probability plots, Kolmogorov–Smirnov tests, and evaluation of skewness and kurtosis.

Comparisons across diagnostic categories (unstable angina, STEMI, NSTEMI) used chi-square tests for categorical variables, one-way ANOVA for normally distributed continuous variables, and Kruskal–Wallis tests for non-normally distributed continuous variables. Sex-based comparisons within each diagnostic group were performed using chi-square or Mann–Whitney U tests, as appropriate.

Logistic regression was used for binary outcomes, multinomial logistic regression for chest pain type and final diagnosis, and linear regression for continuous outcomes, including door-to-ECG time and length of stay. Sex was the primary exposure, with male as the reference category in all models. Model 1 included sex only. Model 2 was adjusted for age, hypertension, smoking, obesity, diabetes mellitus, dyslipidemia, sedentary lifestyle, and family history of premature coronary artery disease. Door-to-ECG time was additionally analyzed as a binary outcome (>10 minutes), corresponding to the recommended diagnostic target, given its right-skewed distribution. Time variables were analyzed in minutes. Chest pain categories with frequencies too low for stable estimation were retained for descriptive reporting and not entered as separate contrasts.

Analyses were conducted for the entire cohort and stratified by diagnosis. Models additionally adjusted for final diagnosis were fitted where appropriate to account for diagnostic case mix. Supplemental subgroup comparisons were considered exploratory, and their p-values were corrected for multiple comparisons using the Benjamini– Hochberg false discovery rate applied across all supplemental comparisons.

Sensitivity analyses included additional adjustment for final diagnosis, exclusion of the 2020–2021 period, sex-by-diagnosis interaction terms, and E-value quantification of potential unmeasured confounding for the main adjusted associations. To account for the clustered structure of the data, the main models were re-estimated with standard errors clustered by state.

Statistical significance was defined as a two-sided p value <0.05. All analyses were performed using R version 4.3.1 within a Jupyter environment (version 7.0.6).

## Results

### Overall Cohort

A total of 43,691 patients with acute chest pain attended by EMS between 2020 and 2024 were included, with 7,171 (16.4%) having acute coronary syndromes (ACS) (Figure 1). For the ACS cohort, the median age was 63 years [IQR 20]. Men accounted for 68.2% (n=4,894) of the cohort. Final diagnoses were unstable angina in 3,348 patients (46.7%), STEMI in 1,345 (18.8%), and NSTEMI in 2,478 (34.6%). Compared to men, women were older (67 [20] vs 61 [19] years) and smoked less frequently, while having more frequent hypertension, diabetes, and sedentary lifestyle. Chest pain was classified as definitely ischemic more often in men, whereas women more often had presentations classified as probably or possibly ischemic and as anginal equivalents. During hospitalization, women had a lower recorded frequency of cardiac catheterization than men **(Table 1).**Diagnosis-specific sex-stratified baseline characteristics are shown in **Supplemental Tables 1–3**.

**Figure 1.**
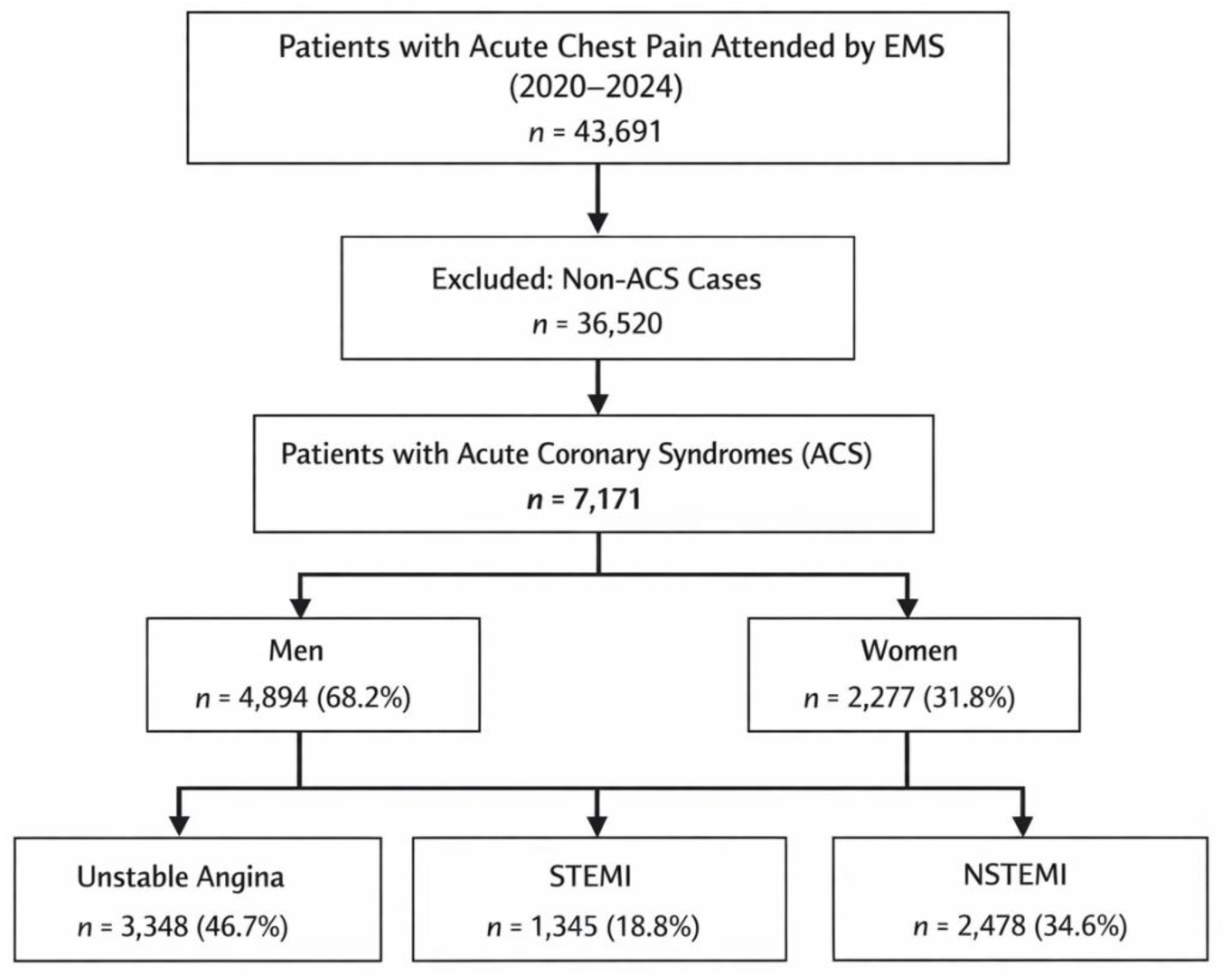
Strobe flow diagram.

**Table 1.** Baseline characteristics, management, and in-hospital outcomes stratified by sex.

| Characteristics | Overall<br>(n=7,171) | Female<br>(n=2,277) | Male (n=4,894) | p-value |
| --- | --- | --- | --- | --- |
| Age (median [IQR]) | 63.0 [20.0] | 67.0 [20.0] | 61.0 [19.0] | <0.001 |
| Cardiovascular Risk Factors |  |  |  |  |
| Hypertension (n, %) | 5446 (76.0) | 1812 (79.6) | 3634 (74.3) | <0.001 |
| Smoking (n, %) | 1310 (18.3) | 358 (15.7) | 952 (19.5) | <0.001 |
| Obesity (n, %) | 1714 (23.9) | 519 (22.8) | 1195 (24.4) | 0.147 |
| Diabetes (n, %) | 2778 (38.7) | 955 (41.9) | 1823 (37.3) | 0.001 |
| Dyslipidemia (n, %) | 3375 (47.1) | 1042 (45.8) | 2333 (47.7) | 0.150 |
| Sedentarism (n, %) | 2214 (30.9) | 745 (32.7) | 1469 (30.0) | 0.030 |
| Family history of CAD (n, %) | 1388 (19.4) | 436 (19.1) | 952 (19.5) | 0.769 |
| Type of Chest Pain (%) |  |  |  |  |
| Definitely ischemic | 1855 (25.9) | 466 (20.5) | 1389 (28.4) | <0.001 |
| Probably ischemic | 3456 (48.2) | 1174 (51.6) | 2282 (46.6) |  |
| Possibly ischemic | 1520 (21.2) | 517 (22.7) | 1003 (20.5) |  |
| Non-ischemic chest pain | 1 (0.0) | 0 (0.0) | 1 (0.0) |  |
| Anginal equivalent | 337 (4.7) | 118 (5.2) | 219 (4.5) |  |
| Evolved myocardial infarction | 2 (0.0) | 2 (0.1) | 0 (0.0) |  |
| Aspirin Administration (%) |  |  |  |  |
| Aspirin 200mg (chewable) | 6576 (91.7) | 2067 (90.8) | 4509 (92.1) | 0.085 |
| Not indicated + Other reason | 418 (5.9) | 153 (6.7) | 265 (5.4) |  |
| Aspirin previous use | 177 (2.5) | 57 (2.5) | 120 (2.5) |  |
| Tirofiban use (n, %) | 69 (1.0) | 15 (0.6) | 54 (1.1) | 0.070 |
| Ticagrelor use (n, %) | 2535 (35.4) | 770 (33.8) | 1765 (36.1) | 0.083 |
| Prasugrel use (n, %) | 745 (10.4) | 150 (6.6) | 595 (12.2) | <0.001 |
| Clopidogrel use (n, %) | 2223 (31.0) | 764 (33.6) | 1459 (29.8) | 0.002 |
| Number of Antiplatelet Agents Administered (%) |  |  |  |  |
| 0 | 169 (2.4) | 52 (2.3) | 117 (2.4) | <0.001 |
| 1 | 2401 (33.5) | 864 (37.9) | 1537 (31.4) |  |
| 2 | 4509 (62.9) | 1326 (58.2) | 3183 (65.0) |  |
| 3 | 92 (1.3) | 35 (1.5) | 57 (1.2) |  |
| Procedures During Hospitalization (%) |  |  |  |  |
| Not performed | 796 (11.1) | 336 (14.8) | 460 (9.4) | <0.001 |
| Not applicable | 425 (5.9) | 196 (8.6) | 229 (4.7) |  |
| Performed | 5949 (83.0) | 1744 (76.6) | 4205 (85.9) |  |
| Non-invasive Stratification (%) |  |  |  |  |
| Not performed | 5626 (78.5) | 1717 (75.4) | 3909 (79.9) | <0.001 |
| Not applicable | 368 (5.1) | 165 (7.2) | 203 (4.1) |  |
| Performed | 1177 (16.4) | 395 (17.3) | 782 (16.0) |  |
| Cardiac Catheterization (n, %) | 5388 (75.1) | 1599 (70.2) | 3789 (77.4) | <0.001 |
| Number of Coronary Arteries Treated (%) |  |  |  |  |
| 1 | 2567 (61.7) | 732 (64.0) | 1835 (60.8) | 0.040 |
| 2 | 1096 (26.3) | 296 (25.9) | 800 (26.5) |  |
| 3 | 499 (12.0) | 115 (10.1) | 384 (12.7) |  |
| Final Diagnosis |  |  |  |  |
| Unstable Angina | 3348 (46.7) | 1197 (52.6) | 2151 (44.0) | <0.001 |
| STEMI | 1345 (18.8) | 329 (14.4) | 1016 (20.8) |  |
| NSTEMI | 2478 (34.6) | 751 (33.0) | 1727 (35.3) |  |
| Outcomes |  |  |  |  |
| In-hospital mortality (n, %) | 179 (2.5) | 50 (2.2) | 129 (2.6) | 0.300 |
| Length of stay in closed unit (ICU/CCU) (days) | 4.0 [4.0] | 4.0 [4.0] | 4.0 [4.0] | 0.296 |
| Hospital length of stay (days) | 5.0 [4.0] | 4.0 [4.0] | 5.0 [4.0] | 0.092 |
| Door-to-needle time (minutes) | 25.0 [10.0] | 23.5 [13.3] | 26.0 [9.0] | 0.323 |
| Door-to-ECG time (minutes) | 4.0 [8.0] | 5.0 [7.0] | 4.0 [8.0] | <0.001 |
| Door-to-guidewire time (minutes) | 72.0 [31.5] | 76.0 [30.0] | 70.0 [31.0] | 0.040 |

### Chest Pain Characteristics and Time Metrics

Across the cohort, chest pain was classified as definitely ischemic in 25.9%, probably ischemic in 48.2%, possibly ischemic in 21.2%, and anginal equivalents in 4.7%, with non-ischemic chest pain and evolved myocardial infarction being rare. Definitely ischemic chest pain predominated in STEMI (73.2%) but was uncommon in unstable angina (11.3%). Probably ischemic and possibly ischemic patterns were more common in patients with angina and NSTEMI (**Table 2**).

**Table 2.**
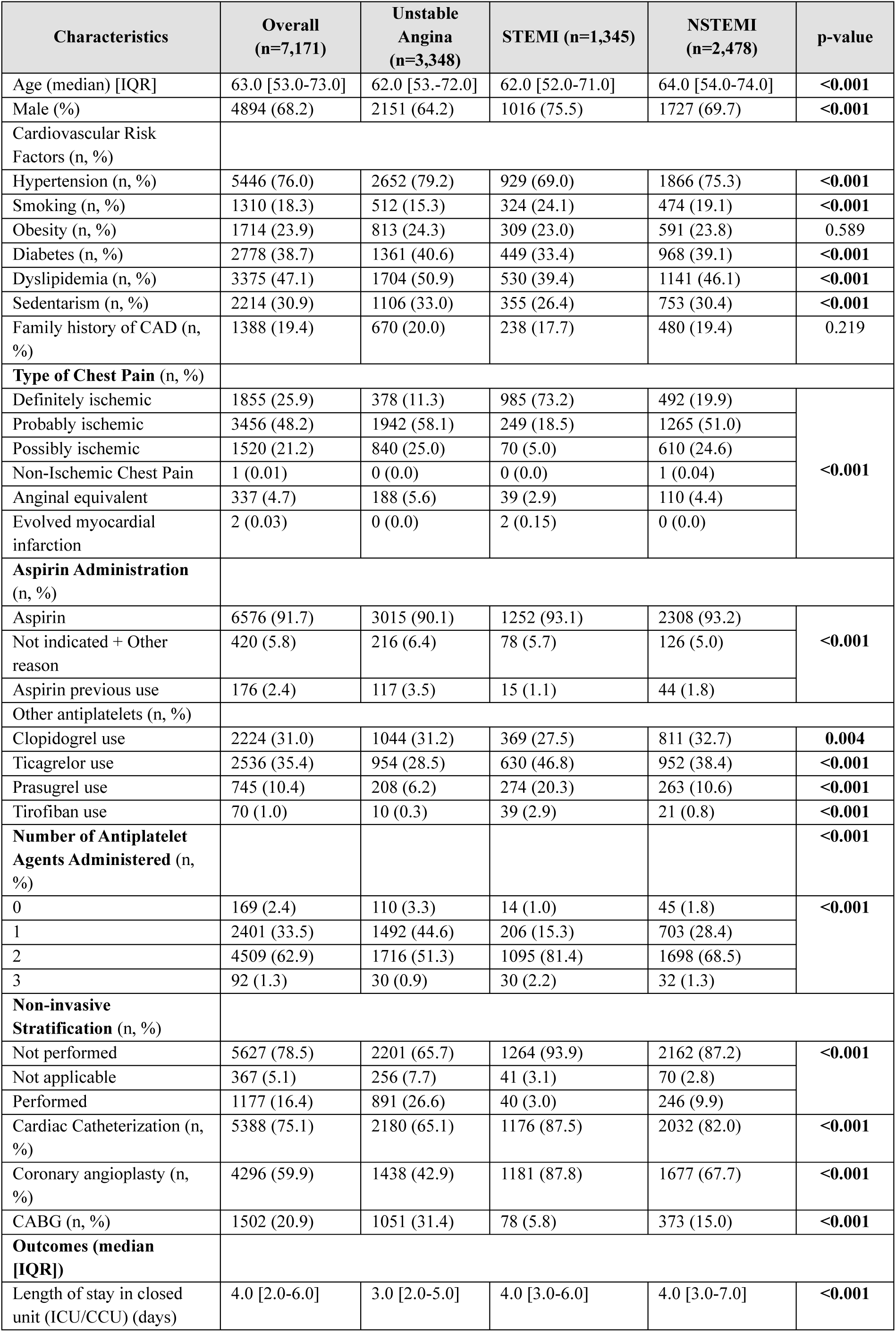
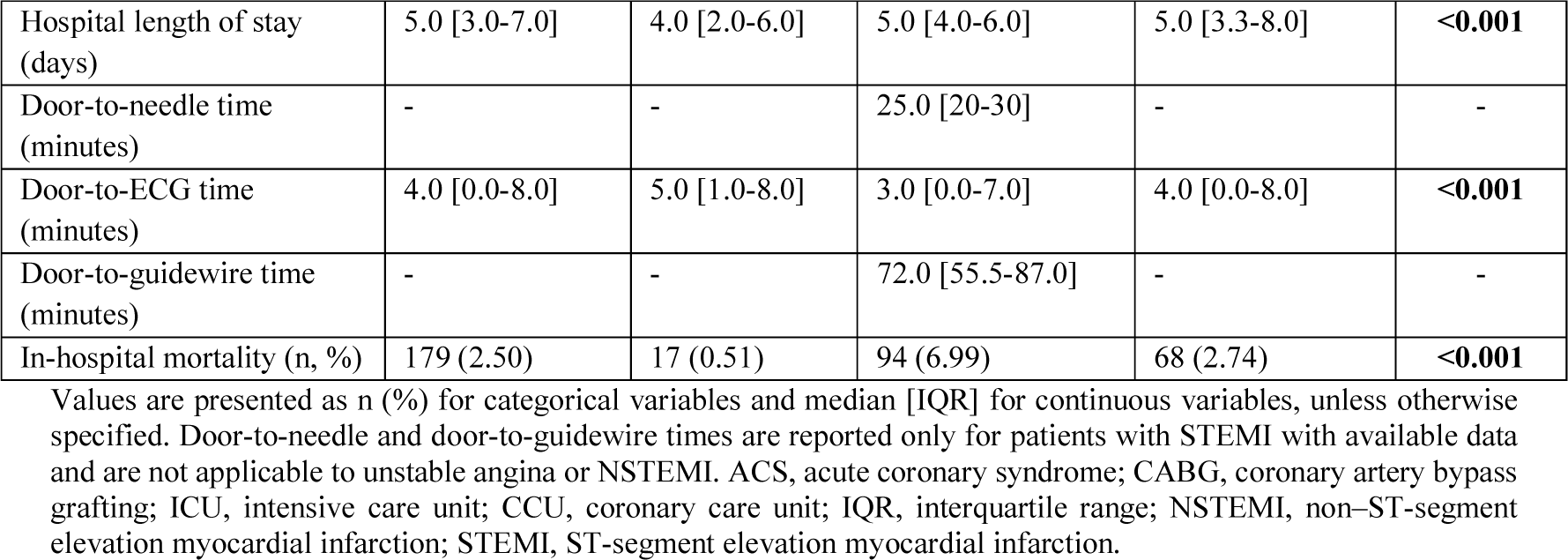
Baseline characteristics, management, and in-hospital outcomes stratified by final ACS diagnosis.

The overall median door-to-ECG time was 4 minutes [IQR 8], and differed across diagnostic categories (p=0.009), with shorter times in STEMI than in unstable angina and NSTEMI (**Table 2)**. In sex-stratified analyses, door-to-ECG time was slightly shorter in men than in women (4 [IQR 8] vs. 5 [IQR 7] minutes; **Table 1**). Coronary angioplasty was recorded in 87.8% of patients with STEMI and was less frequent in NSTEMI (67.7%) and unstable angina (42.9%). Non-invasive stratification was more common in unstable angina and NSTEMI.

Aspirin was administered to 93.1% of patients with STEMI. Among patients with STEMI, 46.8% received ticagrelor, 27.5% clopidogrel, 20.3% prasugrel, and 2.9% tirofiban. Among patients with NSTEMI, 93.2%, 38.4%, 32.7%, 10.6% and 0.8% received aspirin, ticagrelor, clopidogrel, prasugrel, and tirofiban, respectively **(Figure 2)**.

**Figure 2.**
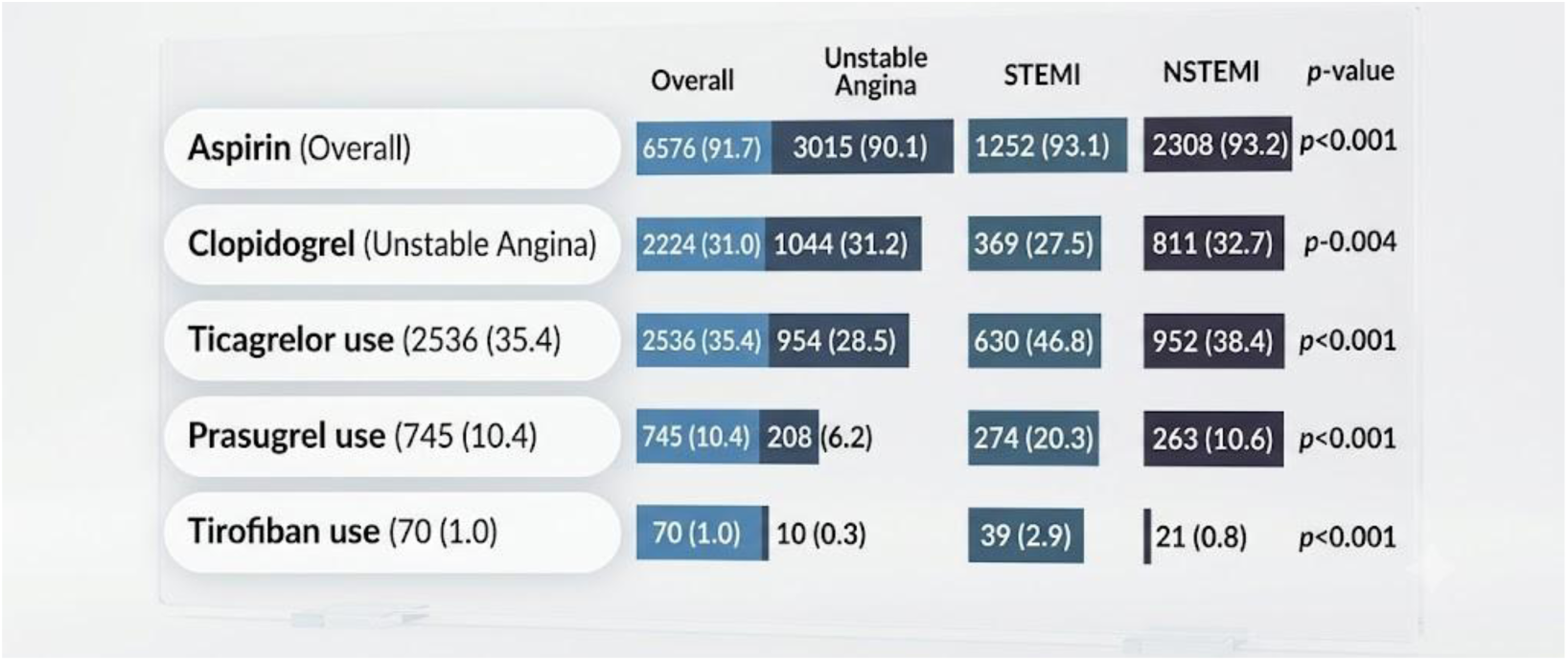
Treatment characteristics according to acute coronary syndrome presentation. Distribution of antiplatelet therapies in the overall study population and according to clinical presentation: unstable angina, ST-segment elevation myocardial infarction (STEMI), and non-ST-segment elevation myocardial infarction (NSTEMI). Data are presented as number of patients (percentage). *P*-values represent comparisons across clinical presentation groups.

### Sex Differences Within Diagnostic Categories

#### Unstable Angina

In the unstable angina subgroup (n=3,348), 1,197 (35.8%) were women and 2,151 (64.2%) were men. Women were older than men (66 [19] vs. 61 [18] years) and had a different cardiovascular risk profile, with more frequent sedentary lifestyle (35.8% vs. 31.5%) and less frequent smoking (13.6% vs. 16.3%) and dyslipidemia (48.0% vs. 52.5%). Hypertension was numerically more frequent among women (81.0% vs. 78.2%), whereas diabetes was similar between groups (41.7% vs. 40.0%). Door-to-ECG time was slightly longer in women (5.0 [1.0–8.0] vs. 4.0 [0.5–8.0] minutes; p=0.011), while closed-unit length of stay was similar.

#### STEMI

In STEMI (n=1,345), 329 (24.5%) were women and 1,016 (75.5%) were men. Women were older than men (68 [19] vs 60 [18] years) and more likely to have diabetes. Hypertension, dyslipidemia, and smoking were prevalent in both sexes. Cardiac catheterization and coronary angioplasty were frequent in both groups, although recorded coronary angioplasty was slightly less frequent in women than in men (84.2% vs. 89.0%). Aspirin administration was high in both sexes. In unadjusted analysis, door-to-ECG time did not differ significantly by sex (p=0.051).

#### NSTEMI

In NSTEMI (n=2,478), 751 (30.3%) were women and 1,727 (69.7%) were men. Women were older than men (69 [20] vs 62 [20] years) and more likely to have hypertension and diabetes, whereas men more often had smoking, and obesity. Coronary angioplasty was less common in women (61.1% vs 70.5% in men), whereas rates of cardiac catheterization were high in both sexes. ICU/CCU length of stay and total hospital length of stay did not differ significantly by sex. Door-to-ECG time was longer in women in unadjusted analyses (p=0.002).

#### Pharmacologic Management

Across the entire cohort, 91.7% of patients received aspirin, 62.9% received dual antiplatelet therapy, 31.0% received clopidogrel, and 35.4% received ticagrelor. Ticagrelor and prasugrel use was highest in STEMI and NSTEMI, whereas clopidogrel use was common across all ACS presentations.

Aspirin use was high in both sexes. Clopidogrel was slightly more commonly used in women (33.6% vs 29.8% in men, p=0.002), whereas prasugrel was more commonly used in men (12.2% vs 6.6% in women, p<0.001). Men more often received two antiplatelet agents (65.0% vs 58.2% in women).

#### Outcomes and times

Closed-unit length of stay was 4.0 days (IQR 2.0-6.0), higher in STEMI (4.0 [3.0-6.0]) and NSTEMI (4.0 [3.0-7.0]) than in UA (3.0 [2.0-5.0], p<0.001), with similar lengths of stay in both sexes. Similarly, hospital length of stay was 5.0 (IQR 3.0-7.0) days overall, higher in STEMI (5.0 [4.0-6.0]) and NSTEMI (5.0 [IQR 3.3-8.0]) than in UA (4.0 [IQR 2.0-6.0]), p <0.001, with no sex differences.

Overall, door-to-ECG time was 4.0 [0.0–8.0] minutes and differed across final ACS diagnoses, with lower values in STEMI (3.0 [0.0–7.0]) and NSTEMI (4.0 [0.0–8.0]) than in unstable angina (5.0 [1.0–8.0], p<0.001). Men had shorter door-to-ECG time than women (4 [0–8] vs. 5 [1–8] minutes), and the proportion beyond the 10-minute target was 11.8% in men and 15.1% in women (**Figure 3**). Diagnosis-specific sex-stratified door-to-ECG distributions are shown for unstable angina and STEMI in **Supplemental Figure 1 and for NSTEMI in Supplemental Figure 2.**

**Figure 3.**
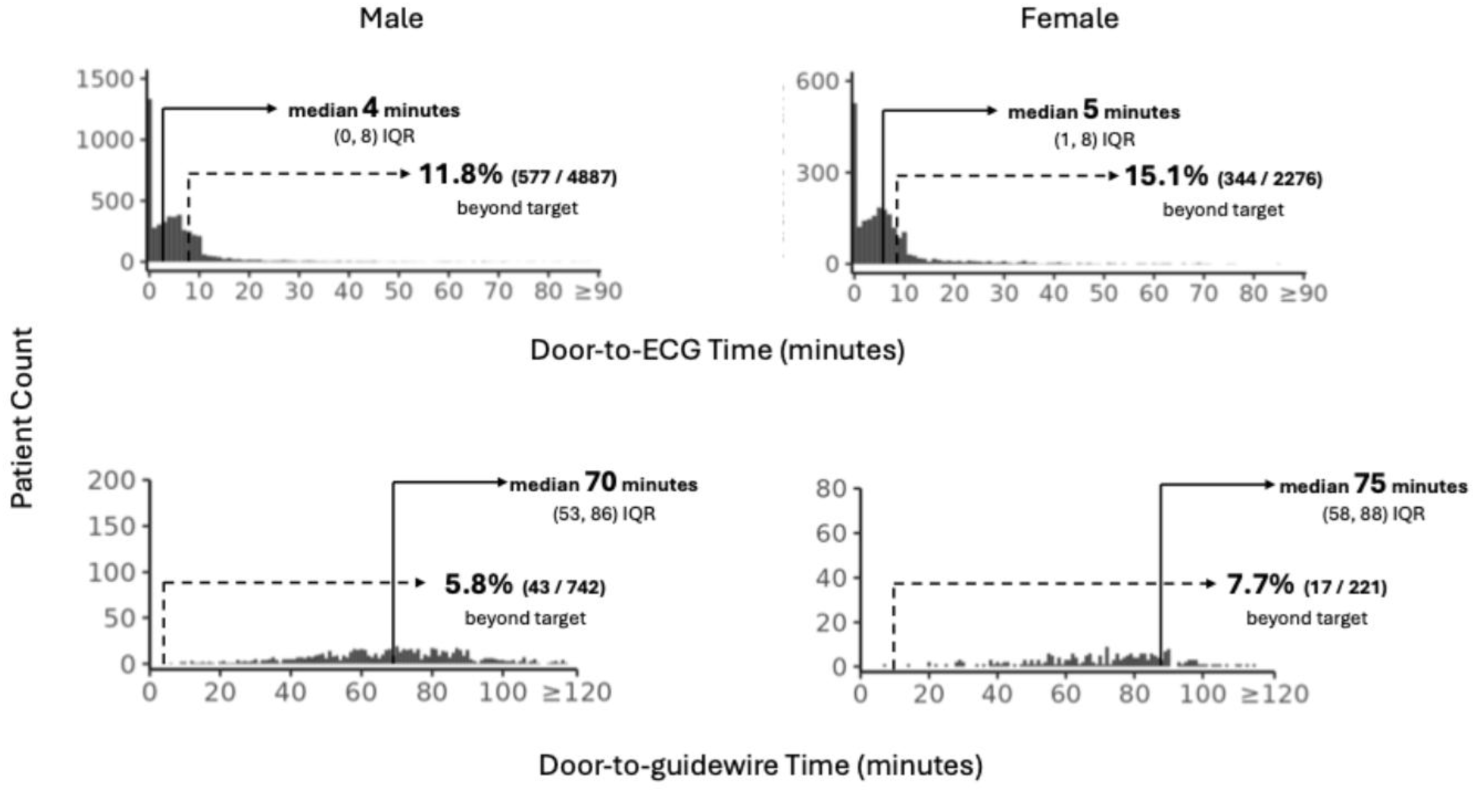
Door-to-ECG and door-to-guidewire times according to sex. Door-to-ECG time was assessed among patients with available data (n = 7,163), whereas door-to-guidewire time was assessed among patients with ST-segment elevation myocardial infarction (STEMI) and available nonnegative values (n = 963). Values exceeding the recommended targets were defined as a door-to-ECG time >10 minutes and a door-to-guidewire time >120 minutes. Data are presented as median [interquartile range (IQR)] or n/N (%), as appropriate.

Among STEMI patients, door-to-needle time was 25.0 minutes (IQR 20.0-30.0) and door-to-guidewire time was 72.0 minutes [55.5-87.0]; door-to-guidewire time was available for 963 STEMI patients (71.6%). Door-to-guidewire time was nominally longer in women than in men (75 [58–88] vs 70 [53–86] minutes), a difference that did not persist after correction for multiple comparisons. In-hospital mortality was 2.5% overall, based on 179 deaths, and was higher in STEMI (6.99%) than in NSTEMI (2.74%) and unstable angina (0.51%) (p<0.001). Mortality did not differ between men (2.6%) and women (2.2%) in unadjusted analysis (p=0.30).

### Multivariable Regression Analyses

#### Overall Cohort

After adjustment for age and cardiovascular risk factors, female sex was associated with higher odds of chest pain classified as probably ischemic or possibly ischemic, relative to definitely ischemic chest pain (probably vs. definitely ischemic, OR 1.51 [95% CI 1.33– 1.72]; possibly vs. definitely ischemic, OR 1.59 [1.37–1.86]; anginal equivalent vs definitely ischemic, OR 1.34 [1.04–1.73]; **Table 3A**. Women were less likely than men to be classified with STEMI or NSTEMI relative to unstable angina (STEMI vs unstable angina, OR 0.59 [0.51–0.68]; NSTEMI vs unstable angina, OR 0.74 [0.66–0.83]; **Table 3A**).

**Table 3A.** Logistic and multinomial logistic regression models comparing sexes (female n=2,277 vs. male n=4,894) for categorical outcomes, considering the total population (n=7,171).

| Outcomes | Observed outcome data, n | Model 1 OR (95% CI); p-value* | Model 2 OR (95% CI); p-value* |
| --- | --- | --- | --- |
| Antiplatelet agents | 7,150 | 0.81 (0.69–0.95); p=0.009 | 0.79 (0.67–0.93); p=0.004 |
| Type of chest pain | 7,123 |  |  |
| Definitely ischemic |  | Reference | Reference |
| Probably ischemic |  | 1.53 (1.35–1.74); p<0.001 | 1.51 (1.33–1.72); p<0.001 |
| Possibly ischemic |  | 1.54 (1.32–1.78); p<0.001 | 1.59 (1.37–1.86); p<0.001 |
| Anginal equivalent |  | 1.60 (1.25–2.05); p<0.001 | 1.34 (1.04–1.73); p=0.025 |
| Aspirin administration | 7,113 |  |  |
| Aspirin administered |  | Reference | Reference |
| Not indicated/other reason |  | 1.26 (1.03–1.55); p=0.028 | 1.23 (1.00–1.52); p=0.049 |
| Previous aspirin use |  | 1.03 (0.75–1.43); p=0.839 | 0.97 (0.70–1.35); p=0.854 |
| Final diagnosis | 7,171 |  |  |
| Unstable angina |  | Reference | Reference |
| STEMI |  | 0.58 (0.50–0.67); p<0.001 | 0.59 (0.51–0.68); p<0.001 |
| NSTEMI |  | 0.78 (0.70–0.87); p<0.001 | 0.74 (0.66–0.83); p<0.001 |

Door-to-ECG time was longer in women in the unadjusted model (β=1.53 minutes [0.24– 2.82]; p=0.020) but the difference was no longer significant after adjustment (β=1.04 minutes [−0.27 to 2.36]; p=0.119; **Table 3B**). When door-to-ECG time was modeled as exceeding the 10-minute target, the sex difference was not supported in the wild cluster bootstrap sensitivity analysis by state. No sex differences were observed in hospital or closed-unit length of stay (**Table 3B**). The lower adjusted odds of the recorded antiplatelet regimen in women (OR 0.79 [0.67–0.93]) did not persist after additional adjustment for final diagnosis (OR 0.85 [0.73–1.01]; p=0.058; **Supplemental Table 4**).

**Table 3B.** Linear regression comparing sexes (female n=2,277 vs. male n=4,894) for continuous outcomes, considering the total population (n=7,171).

| Outcomes | n analyzed | Model 1 $\beta$ (95% CI); p-value* | Model 2 $\beta$ (95% CI); p-value* |
| --- | --- | --- | --- |
| Door-to-ECG time, minutes | 7,163 | 1.53 (0.24 to 2.82); p=0.020 | 1.04 (–0.27 to 2.36); p=0.119 |
| Length of stay in closed unit (ICU/CCU), days | 6,096 | –0.32 (–1.23 to 0.59); p=0.490 | –0.678 (–1.61 to 0.25); p=0.154 |
| Hospital length of stay, days | 7,171 | 0.03 (–0.38 to 0.44); p=0.881 | –0.29 (–0.70 to 0.12); p=0.170 |

In-hospital mortality did not differ by sex in the unadjusted model (OR 0.83 [0.60–1.15]; p=0.267). In the model adjusted for age and cardiovascular risk factors, the point estimate was lower in women (OR 0.59 [0.42–0.83]); this estimate was sensitive to model specification (see Sensitivity Analyses). Overall, there was no evidence of excess short-term mortality in women.

#### Sensitivity Analyses

The main associations for ischemic chest pain classification and final diagnosis were consistent across temporal sensitivity analyses. After adjustment for the attendance period (2020–2021 vs. 2022–2024), the estimates remained similar for probably ischemic chest pain (OR 1.51 [1.33–1.72]), possibly ischemic chest pain (OR 1.60 [1.37–1.86]), STEMI versus unstable angina (OR 0.59 [0.51–0.68]), and NSTEMI versus unstable angina (OR 0.74 [0.66–0.83]).

Excluding the early pandemic years (2020–2021), these associations remained significant for probably ischemic chest pain (OR 1.39 [1.19–1.61]), possibly ischemic chest pain (OR 1.51 [1.27–1.80]), STEMI versus unstable angina (OR 0.59 [0.49–0.70]), and NSTEMI versus unstable angina (OR 0.75 [0.65–0.85]). In contrast, anginal equivalent symptoms showed the greatest attenuation among the chest pain categories: the association was attenuated to borderline significance after exclusion of the early pandemic years (OR 1.33 [1.00–1.78]; p=0.052) and did not remain statistically significant in the state-cluster sensitivity analysis using wild cluster bootstrap. In that analysis, the associations for probably ischemic and possibly ischemic chest pain, as well as for STEMI and NSTEMI versus unstable angina, remained statistically significant, whereas the diagnosis-adjusted association for the recorded antiplatelet regimen was no longer statistically significant **(Supplemental Table 4).**

For in-hospital mortality, the lower adjusted odds in women were not consistent across specifications: the estimate moved toward the null when excluding the pandemic years (OR 0.68 [0.45–1.02]; p=0.062) and when accounting for clustering by state, and the E-value at the confidence-interval limit was 1.28.

E-values at the confidence-interval limit were 1.99 for probably ischemic, 2.08 for possibly ischemic, 1.24 for anginal equivalent, 2.30 for STEMI vs unstable angina, and 1.72 for NSTEMI vs unstable angina. No sex-by-diagnosis interaction was observed for chest pain category (p=0.211) or in-hospital mortality (p=0.405). Among 125 supplemental comparisons, 44 had nominal p-values <0.05, of which 28 remained significant after Benjamini–Hochberg correction

#### STEMI Subgroup

In exploratory analyses of the STEMI subgroup, no significant differences were observed between women and men in antiplatelet use or in hospital or closed-unit length of stay. Door-to-ECG time did not differ significantly by sex (p=0.051). Door-to-guidewire time was nominally longer in women (p=0.040) but this difference did not persist after correction for multiple testing. Similarly, no significant difference was found for door-to-needle time.

## Discussion

In this large, multicenter EMS-based study of 7,171 Brazilian patients with acute chest pain, we observed sex-related differences in ACS presentation, symptom classification, and diagnostic categorization. Women were older, more frequently presented with chest pain classified as less definitely ischemic and were less often classified as having STEMI or NSTEMI relative to unstable angina. In contrast, sex-related differences in downstream management were less pronounced. The modest difference in door-to-ECG time observed in unadjusted analyses was no longer evident after adjustment for age and cardiovascular risk factors and was not statistically significant in the threshold-based sensitivity analysis. Likewise, the association between sex and the recorded antiplatelet regimen was attenuated after additional adjustment for final diagnosis. Overall, the findings suggest that sex-related differences in this cohort were concentrated in the early stages of clinical assessment and diagnostic categorization rather than in subsequent treatment or short-term in-hospital outcomes.

These results are consistent with prior reports from high-income countries, which have highlighted older age at presentation, atypical symptoms, and diagnostic delays. ^9^ The disparities in risk profiles are consistent with the Global Registry of Acute Coronary Events from 14 countries, where women with ACS were older and had higher rates of hypertension (70% versus 55%) and diabetes (29% versus 22%).^10,11^ Despite these differences, women in our cohort had crude in-hospital mortality like that of men, contrasting with previous data.^12^ After adjustment for age and cardiovascular risk factors, the point estimate for in-hospital mortality was lower in women. However, this association varied across temporal and clustering sensitivity analyses, and the registry did not include several markers of disease severity known to influence short-term mortality. Accordingly, our findings do not support an excess short-term mortality among women in this network but should not be interpreted as evidence of a sex-related survival advantage. A national chest pain protocol implemented across this hospital network may have contributed to standardized care delivery, however the absence of a non-protocolized comparator group precludes attributing these findings directly to the protocol.

Sex-related differences in this cohort appeared to concentrate in symptom classification and diagnostic categorization rather than subsequent therapeutic decisions. Women had higher odds of chest pain classified as probably or possibly ischemic and lower odds of STEMI or NSTEMI relative to unstable angina, suggesting that these patterns should be interpreted along the same diagnostic pathway rather than as isolated observations. Older age, greater comorbidity burden, and less typical symptom patterns may have contributed to classification as less definitely ischemic chest pain.

The associations for probably ischemic and possibly ischemic chest pain were among the most consistent results, persisting in temporal sensitivity analyses, remaining statistically significant in the state-cluster wild bootstrap analysis, and showing E-values at the confidence-interval limit close to 2.0. Anginal equivalent symptoms showed greater attenuation, reaching borderline significance after exclusion of the early pandemic years and losing statistical significance in the state-cluster wild bootstrap analysis. The associations for final diagnosis also showed E-values at the confidence-interval limit of 2.30 for STEMI and 1.72 for NSTEMI relative to unstable angina, indicating that unmeasured confounding of at least moderate magnitude would be required to fully explain the observed associations involving symptom classification and diagnostic categorization.

Biological differences should also be considered when interpreting sex-related differences in ACS presentation. Women presenting with ACS are twice as likely to present with myocardial infarction in nonobstructive coronary arteries (MINOCA) as men are.^13^ In addition, 87% to 95% of spontaneous coronary artery dissection cases occur in women^14^ and plaque erosion is more common in women than in men. ^15^ These mechanisms provide biologically plausible explanations for differences in clinical presentation and diagnostic categorization; however, the present registry did not systematically adjudicate MINOCA, spontaneous coronary artery dissection, or plaque erosion.

Women with ACS have historically experienced lower rates of evidence-based therapies, including invasive angiography, revascularization, and dual antiplatelet therapy, partly due to under-recognition of risk and the perception of lower cardiovascular risk among women. ^16^ ^17^ ^18^ By comparison, aspirin use and the recorded antiplatelet regimen were frequent in both sexes in our cohort. Although women had lower adjusted odds of receiving the recorded antiplatelet regimen, this association was attenuated after adjustment for final diagnosis and was no longer statistically significant in the state-cluster sensitivity analysis, suggesting that diagnostic case mix, rather than sex itself, may explain the observed difference. Among patients with STEMI, PCI without documented fibrinolysis was the predominant reperfusion strategy, and coronary angioplasty was performed in a high proportion of both women and men. Procedure-level comparisons by sex were limited by differential availability of procedural records and were therefore not interpreted as treatment disparities. Likewise, no significant sex differences were observed in adjusted length of stay, and crude in-hospital mortality was similar between sexes. Overall, these observations suggest that, among patients with confirmed ACS, the measurable treatment differences were limited, directing attention primarily to earlier stages of clinical assessment and diagnostic classification rather than to subsequent management.

Protocol-driven systems of care have been associated with reduced variability in clinical decision-making and improved adherence to guideline-recommended therapies.^19^ In our network, all participating EMS systems operated under a unified chest pain protocol with cardiologist oversight, which may have contributed to the high rates of aspirin administration, dual antiplatelet therapy, and invasive management observed in both sexes. However, because all participating sites used the protocol, this study cannot determine whether the protocol itself reduced sex-related disparities.

Another aspect that warrants consideration is the distinction between Brazil’s private and public healthcare sectors. The hospitals included in this study are part of the largest private hospital network in the country and generally have greater availability of cardiovascular specialists, catheterization laboratories, telecardiology support, and rapid diagnostic resources than many public institutions.^20^ Consequently, our findings may not be fully generalizable to the broader Brazilian healthcare system. Future studies should evaluate whether similar sex-related patterns are present within the public healthcare system and whether protocolized approaches can similarly mitigate disparities in ACS recognition and management.

Women had longer door-to-ECG times in unadjusted analyses, but the magnitude was modest and the adjusted difference was attenuated after accounting for age and cardiovascular risk factors. This suggests that part of the crude difference in ECG timing may reflect the older age and greater clinical complexity of women, rather than an independent effect of sex. Clinically, this distinction matters: the quality-improvement target may not be women as a homogeneous group, but rather older, more comorbid women with less typical or less definitely ischemic presentations. Timely ECG acquisition remains essential in suspected ACS, particularly in STEMI, where it triggers activation of reperfusion pathways; in this cohort, sex-related differences in ECG timing were not sustained after adjustment or in the threshold-based analysis. Among patients with STEMI and available door-to-guidewire data, median door-to-guidewire time was nominally longer in women than in men, however this measure unavailable for a substantial proportion of STEMI patients, the proportions beyond the 120-minute target were similar, and the difference did not persist after correction for multiple comparisons, This finding should therefore be interpreted as descriptive. Symptom interpretation and triage processes remain important targets for quality improvement initiatives.

The adjusted analyses further align with this interpretation. Women were more likely than men to be classified as having probably or possibly ischemic chest pain, rather than definitely ischemic chest pain, highlighting persistent differences in how presentations were categorized. Because this cohort was restricted to patients with ACS confirmed as the final diagnosis, these differences in symptom classification were observed after patients had already crossed the threshold for confirmed ACS. Thus, the measured difference may represent the portion of sex-related presentation patterns that remains visible after diagnostic confirmation; the study, however, cannot directly assess under-recognition before cohort entry. These findings are consistent with prior literature demonstrating that ACS in women frequently manifests through less common symptoms, including dyspnea, fatigue, nausea, epigastric discomfort, or generalized weakness rather than classic substernal chest pain.^9^ Recognition of these presentations remains critical for reducing diagnostic delays.

Future research should focus on evaluating sex-specific risk stratification tools, artificial intelligence-assisted triage systems, and educational interventions designed to improve recognition of ACS among women in prehospital settings. In addition, studies examining long-term outcomes, recurrent cardiovascular events, and quality-of-life measures would provide a more comprehensive understanding of the impact of sex differences across the entire continuum of ACS care.

From a clinical and policy perspective, these findings underscore the need for sex-sensitive triage algorithms that explicitly account for atypical ACS presentations, particularly in older women with multiple risk factors. Protocolized ECG timing and automated prompts for ECG acquisition in all patients with chest discomfort or less typical ACS-compatible presentations—regardless of sex—may help close the diagnostic gap. Educational initiatives addressing implicit bias and sex-specific ACS patterns for EMS personnel, combined with monitoring of sex-stratified quality metrics, could further promote equity in care.

Overall, our findings indicate that, within a protocolized private EMS-hospital network, the main sex-related differences were concentrated in symptom classification and diagnostic categorization rather than in adjusted length of stay or crude short-term mortality. The data are consistent with a model in which women’s older age, comorbidity burden, and less definitely ischemic presentation are associated with diagnostic classification, while downstream treatment markers appeared less consistently sex-differentiated after accounting for diagnostic case mix. Procedure-level comparisons, however, were limited by differential record availability. Opportunities remain to improve the earliest phases of care, particularly symptom interpretation among women presenting with suspected ACS.

### Strengths and Limitations

Strengths of this study include the large sample size, a multicenter design spanning 14 Brazilian states, and the linkage of EMS and hospital data. The stratified analyses by diagnosis (unstable angina, STEMI, and NSTEMI) allowed us to explore heterogeneity in sex-based differences across the ACS spectrum. The main associations were examined across temporal sensitivity analyses, state-cluster wild bootstrap sensitivity analyses, sex-by-diagnosis interaction testing, false discovery rate correction for supplemental comparisons, and E-value quantification of potential unmeasured confounding.

Limitations include the retrospective design and potential residual confounding and misclassification. The cohort comprised patients with ACS confirmed as the final diagnosis; therefore, the study evaluated sex-related differences in classification and management within patients already recognized as having ACS, rather than differences in recognition at first contact among all patients presenting with chest pain. If under-recognition of ACS in women occurs before diagnostic confirmation, it would occur prior to cohort entry and could not be directly measured in this design. Rare categories within some variables also produced unstable regression estimates and should be interpreted with caution. Supplemental subgroup comparisons were exploratory, and several nominal associations did not persist after FDR correction. The lower adjusted odds of in-hospital mortality in women emerged only after adjustment and were not consistent across temporal and clustering specifications; in addition, the registry did not capture measures of clinical severity, such as Killip class, ventricular function, biomarkers, or renal function, that are known to influence mortality. This finding should therefore be regarded as hypothesis-generating and is not interpreted as a sex-related difference in survival. Procedure-level differences by sex, including catheterization and angioplasty, were affected by substantial and sex-differential missingness in procedural records, with higher missingness among women, and were therefore not interpreted as definitive treatment disparities. Finally, long-term outcomes beyond hospital discharge were not available.

### Conclusions

In a large private EMS-hospital network in Brazil, women with confirmed ACS were older, more frequently had chest pain classified as less definitely ischemic, and were less often classified as having STEMI or NSTEMI relative to unstable angina. Door-to-ECG time and use of antiplatelet were similar between sexes after adjustment, and no excess short-term mortality was observed among women. Within this protocolized network, sex-related differences in ACS care appeared to be concentrated primarily in symptom classification and diagnostic categorization rather than in persistent adjusted differences in treatment markers or short-term outcomes. Sex-sensitive triage strategies should therefore focus on improving the interpretation and classification of less typical presentations, particularly among older and comorbid women.

## Data Availability

The data that support the findings of this study are not publicly available because they contain information derived from an institutional clinical registry. De-identified data may be made available from the corresponding author upon reasonable request, subject to institutional approval and applicable ethical and data protection requirements.

## Acknowledgments

The authors thank the participating emergency medical services and hospital teams across the 14 Brazilian states, as well as the information technology personnel who maintain the EMS electronic medical record systems.

## Supplemental Figure Legends

Supplemental Figure 1. Sex-stratified door-to-ECG time according to acute coronary syndrome presentation. Door-to-ECG time is shown for female and male patients with unstable angina and ST-segment elevation myocardial infarction (STEMI). Values exceeding the recommended target were defined as a door-to-ECG time >10 minutes. Data are presented as median [interquartile range (IQR)] and n/N (%), as appropriate.

Supplemental Figure 2. Sex-stratified door-to-ECG time among patients with non-ST-segment elevation myocardial infarction. Door-to-ECG time is shown for female and male patients with non-ST-segment elevation myocardial infarction (NSTEMI). Values exceeding the recommended target were defined as a door-to-ECG time >10 minutes. Data are presented as median [interquartile range (IQR)] and n/N (%), as appropriate.

